# High interest in Long-Acting Injectable PrEP among Filipina Transfeminine Adults

**DOI:** 10.1101/2022.08.02.22278323

**Authors:** Arjee Javellana Restar, Ma Irene Quilantang, Jeffrey Wickersham, Alex Adia, John Guigayoma, Amiel Nazer Bermudez, Omar Galárraga, Dalmacio Dennis Flores, Susan Cu-Uvin, Jennifer Nazareno, Don Operario, Olivia Sison

## Abstract

Transfeminine adults are impacted by the HIV epidemic in the Philippines, and newly approved modalities of pre-exposure prophylaxis (PrEP), including long-acting injectable (LAI-PrEP), could be beneficial for this group. We utilized secondary data from the #ParaSaAtin survey that sampled Filipina transfeminine adults (n=139) and conducted a series of multivariable logistic regressions with lasso selection to explore factors independently associated with PrEP outcomes, including awareness, discussion with trans friends, and interest in LAI-PrEP. Overall, 53% of Filipina transfeminine respondents were aware of PrEP, 39% had discussed PrEP with their trans friends, and 73% were interested in LAI-PrEP. PrEP awareness was associated with being non-Catholic, having previously been HIV tested, discussing HIV services with a provider, and having high HIV knowledge (p<0.05). Discussing PrEP with friends was associated with older age, having experienced healthcare discrimination due to transgender identity, having HIV tested, and having discussed HIV services with a provider (p<0.05). Interest in LAI-PrEP was associated with living in Central Visayas, having discussed HIV services with a provider, and having discussed HIV services with a sexual partner were associated (p<0.05). Implementing LAI-PrEP in the Philippines requires addressing systemic improvements across personal, interpersonal, social, and structural levels in healthcare access, including efforts to create healthcare settings and environments with providers who are trained and competent in transgender health and can address the social and structural drivers of trans health inequities, including HIV and barriers to LAI-PrEP.

## Introduction

Transfeminine adults are disproportionately impacted by the growing HIV epidemic in the Philippines, where the epidemic has grown exponentially over the last decade [1]. Based on successful findings from a PrEP demonstration study in the Philippines showing clinical effectiveness [2], the Philippines’ Department of Health recently approved Tenofovir/Emtricitabine (TDF/FTC) as daily oral pre-exposure prophylaxes (PrEP) for HIV prevention [3]. Updated national clinical guidelines now recognize PrEP as part of its national essential medicine formulary as of January 2022 [3], and recommend the urgent need to deliver it to populations placed vulnerably to HIV infection, including Filipina transfeminine adults who are bearing the brunt of the epidemic – with HIV prevalence approximately doubling (3.9% in 2020 vs. 1.7% in 2018) within a short timeline [1, 4]. Moreover, with several clinical trials in the pipeline testing new modalities such as long-acting injectable pre-exposure prophylaxes (LAI-PrEP) – including the cabotegravir extended-release injectable suspension formulation that was recently approved by the United States Food and Drug Administration that showed superior efficacy when compared to daily oral PrEP among transfeminine adults [5, 6] – understanding how LAI-PrEP would fit into the lives of transfeminine adults in the Philippines is necessary for future PrEP implementation programs in the county.

The emerging literature of implementation science, as applied within the field of biomedical HIV prevention [7, 8], emphasizes the need for a critical, formative understanding of multiple determinants across the socio-ecological (e.g., personal, social, community, structural) levels that can guide implementation of LAI-PrEP. In a systematic review synthesizing determinants of PrEP implementation [8], frequent barriers included factors across socio-ecological levels: (a) at the personal level - lack of PrEP awareness and knowledge, adherence challenges to daily medication over time, and concerns about side effects and interactions with other medications such as gender-affirming hormones [9-12]; (b) at the interpersonal level - discomfort talking to providers about PrEP, provider’s lack of knowledge or support of PrEP, and having mistrust of providers [13-15]; (c) at the social level -lack of partner support for taking PrEP, and lack of communication about health issues and PrEP among community members [14, 16, 17], including PrEP discussion among trans friends and providers [15]; and (d) at the structural level - having segregated or fragmented health care systems, concern about paying for PrEP including out-of-pocket costs for necessary labs and visit copays, frequency of required clinic visits, stigma and discriminatory practices and policies related to PrEP, HIV, transgender identity, and sexual behavior, as well structural determinants such as poverty, unemployment, unstable housing [14, 16-19]. Moreover, while the Philippines is considered one of the most socially tolerant countries for lesbian, gay, bisexual, and transgender (LGBT) people compared to other neighboring countries in the Southeast Asian region [20], its policies and laws continue to reflect transphobia, sexual conservatism, and highly influenced by pervasive religious fundamentalist hegemony [21-23], with 81% of the country’s population being affiliated with Catholicism [24].

Given transfeminine adults’ vulnerability to HIV infection in the Philippines as well as the multiple barriers and challenges across the socio-ecological levels in implementing PrEP, understanding promising strategies to deliver new PrEP modalities is vital to its success. While some lessons can be derived from previous PrEP studies, advances in injectable PrEP as a delivery modality could pose unique challenges and barriers to implementation of LAI-PrEP among transfeminine adults in the Philippines, which are necessary considerations for LAI-PrEP program designers and providers to optimize engagement across the PrEP continua (i.e., awareness, uptake, adherence, and retention) [25-29]. To our team’s knowledge, there are currently no studies that specifically examine LAI-PrEP among Filipina transfeminine populations [30]. Given the need for formative studies in LAI-PrEP implementation among Filipina transfeminine adults, the purpose of our exploratory study is to examine prevalence and correlates of outcomes along the PrEP continua, particularly awareness, discussion among friends, and interest in taking LAI-PrEP.

## Methods

### Study Sample and Procedures

We utilized the STROBE checklist for reporting cross-sectional study, which can be found in S1 Table.

This is a secondary data analysis of Filipina transfeminine respondents (n=139) from the #ParaSaAtin project, a cross-sectional online survey conducted between June 2018 to May 2019 that examined the social, community, and structural drivers of HIV and factors that impact the utilization of HIV services including PrEP among transfeminine adults in the Philippines. For this analysis, we focused specifically on PrEP awareness, PrEP discussion among trans friends, and interest in LAI-PrEP as outcomes.

Full study procedures are described elsewhere [31]. Briefly, the study used online convenience sampling recruitment strategies, which included deploying the survey using banners, social media advertisements, and online social groups (e.g., Facebook, Twitter) hosted by local HIV community-based organizations and transgender communities in the Philippines. Participants enrolled in the study were: 18 years or older, transfeminine (i.e, have a gender identity along the transfeminine spectrum such as women, trans women, and with sex assigned at birth as male), had condomless sex within the past year, currently living in Metro Manila and Central Visayas (i.e., the two HIV hotspot areas in the country), and demonstrated English and consent comprehension via a brief cognitive screening form. We utilized multiple best practices for online sampling procedures [32], which include: (1) using a captcha box to eliminate out robots as well as, (2) deactivating ballot stuffing to prevent survey takers from taking the survey more than once. The survey lasted between 20 to 25 minutes, and participants received ₱300 ($5.85 USD) for completing the survey.

### Measures

#### Demographic factors

Participants were asked about their age (18 to 24, 25 to 29, 30 to 34, 35 years or more), current living location (Metropolitan Manila, or Central Visayas), highest education attained (high school and below, some college, or college and beyond), past year income (no income / less than ₱10,000, ₱10,000 to less than ₱20,000, ₱20,000 to less than ₱30,000, ₱30,000 or more), religious affiliation (Catholic, non-Catholic (e.g., Protestant, Christian), or non-religious), and sexual orientation (gay/lesbian, bisexual, straight, or another sexual orientation).

#### Social marginalization factors

Social marginalization included measures specific to homelessness, unemployment, and whether they themselves engaged in sex work. We assessed whether participants had a history of homelessness (yes vs. no), were currently unemployed (yes vs. no), or recently within the past 4 months) had engaged in sex work themselves (yes vs. no).

#### Community-level factors

Community-level factors included measures of social cohesion, general and LGBT-specific social participation, and healthcare accessibility. We adapted Lippman and colleagues’ social cohesion scale [33], which is comprised of 9 items with 5-point Likert responses (from strongly agree=1 to strongly disagree=5) that measure participants’ perceptions of trust and connectedness with their communities, including items such as: “You can count on your friends to help you access health services,” and “You can count on your friends to support the use of condoms.” Responses were tested for internal consistency (Cronbach *α*= 0.87), summed, scored continuously, and split at the median to dichotomize into low vs. high social cohesion categories. We then assessed social participation in general and LGBT community activities by adapting Fonner and colleagues’ 4-item social participation scale [34]. Similarly, responses were tested for internal consistency (Cronbach *α*=0.75 for general social participation, and Cronbach *α*=0.76 for LGBT social participation, respectively), summed, scored continuously, and split at the median to dichotomize into low vs. high social participation categories. Lastly, we adapted Haggerty and colleagues’ 6-item healthcare accessibility scale to assess participant’s level of care access (e.g., traveling to their provider’s office, waiting times) [35]. Responses ranged from very poor to excellent, and were then tested for internal consistency, (Cronbach *α*=0.93), summed, scored continuously, and split at the median to dichotomize into poor/fair vs. good/excellent accessibility categories.

#### HIV and other healthcare indicators

Participants were asked about their health care history and experiences, including if they have current health insurance, ever took hormones for gender affirmation, ever had surgery for gender affirmation, or experienced healthcare discrimination due to their transgender identity. We also asked about their healthcare history with HIV services, including if they ever had an HIV test, discussed HIV with their provider or sexual partner, or avoided HIV services due to: cost, distance to/from a healthcare facility, their transgender identity, or healthcare facility’s lack of LGBT anti-discrimination policy. All responses were either yes or no. Additionally, using the International AIDS Questionnaire, we assessed participants’ HIV knowledge. Responses were tested for internal consistency (Cronbach *α*= 0.84), summed, scored continuously, and split at the median to dichotomize into low vs. high HIV knowledge categories [36].

#### PrEP awareness, PrEP discussion among trans friends, and Interest in LAI-PrEP (outcomes)

We asked whether participants are aware of PrEP, have had discussions about PrEP with their trans friends, and if they are interested in taking LAI PrEP. These questions were adapted from Restar and colleague’s PrEP acceptability study [13]. Specifically, all participants were first provided a brief definition of PrEP: “One way to fight HIV is called PrEP, which stands for pre-exposure prophylaxis. PrEP works by giving HIV-negative people HIV drugs to keep them from getting HIV.” To assess for PrEP awareness, we asked participants if they “have heard of HIV-negative people taking HIV medication before sex because they thought it would lower their chances of getting HIV (also known as PrEP)?” We then asked participants if they have had PrEP discussions among their trans friends. Response options for both questions were either yes vs. no. Lastly, we assessed interest in LAI-PrEP by asking, “If the possibility of having a long-lasting injectable drug to prevent HIV (“injectable PrEP”) was available, would you be interested in taking it?” and responses were conservatively recoded and dichotomized to very interested vs. somewhat/not at all interested.

### Data Analysis

We conducted descriptive analyses (frequencies, percentages) to examine the overall distribution of the final analytic sample (n=139) of Filipina transfeminine adults, and stratified per our outcomes (PrEP awareness, PrEP discussion, and interest in LAI-PrEP). We then performed bivariate analyses to examine patterns of our outcomes across our measured exposure variables (e.g., demographics, social marginalization factors, community factors, HIV and other healthcare indicators).

To examine characteristics associated with our outcomes (PrEP awareness, PrEP discussion, and interest in LAI-PrEP), we performed a series of multivariate logistic regression analyses. Given the exploratory nature of this study, we utilized lasso regression procedures to identify the key variables per outcome [37]. All variables that were selected via the lasso procedure were included in the final adjusted model per outcome. Additionally, given the modest sample size, we used nonparametric bootstrapping procedures using 100 iterations to reduce Type 1 error per model and estimate confidence intervals [38]. Alpha for analyses was set to <0.05 a priori, and all analyses were conducted using StataMP version 17.0.

### Ethics

Electronic, written consent forms were obtained from all participants in this study, which provided information about participants’ confidentiality, privacy, and voluntary participation in this study. This study acquired approval from Institutional Review Board of the Brown University Brown University Human Research Protection Program (IRB Protocol, #1802001982).

## Results

### Sample Characteristics

Shown in Table 1, most respondents in the sample were between 25-29 years old (39%), lived in the Metropolitan Manila area (79%), attained college education or beyond (44%), earned less than ₱10,000 or no income in the past year (55%), Catholic (79%), and gay or lesbian (55%). A total of 27% of the sample had ever been homeless, 42% currently unemployed, and 52% had themselves recently engaged in sex work.

**Table 1.**
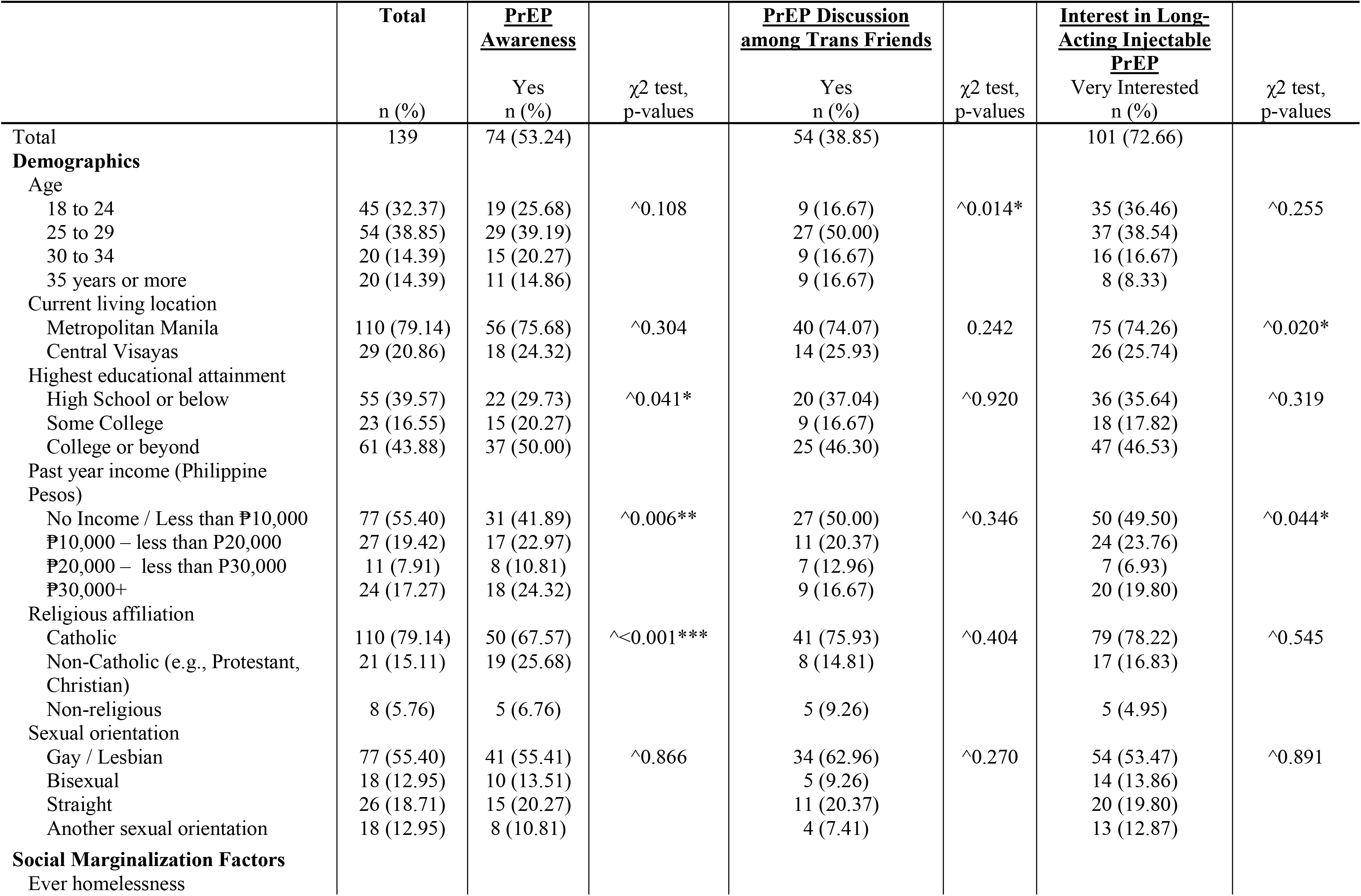
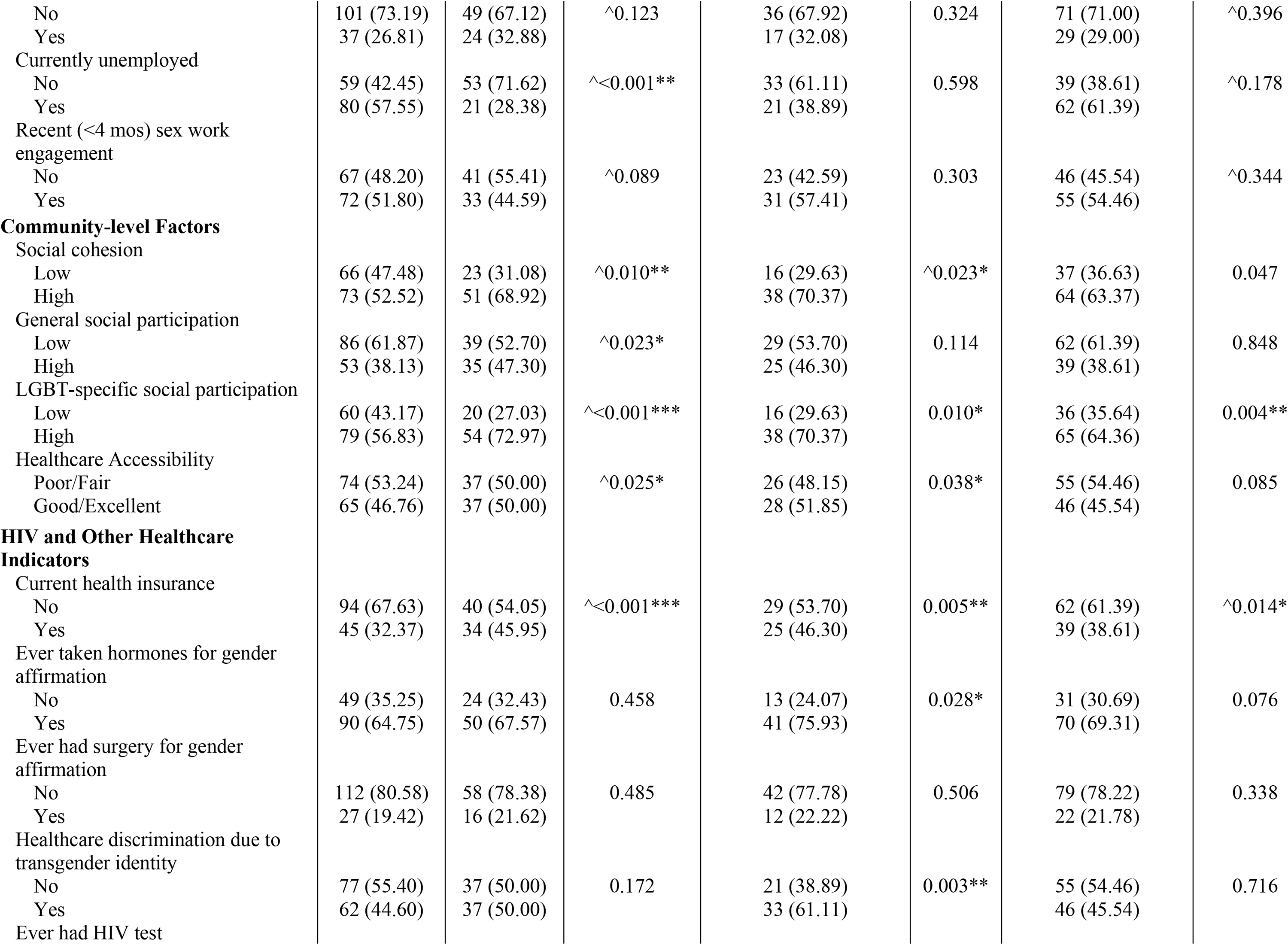
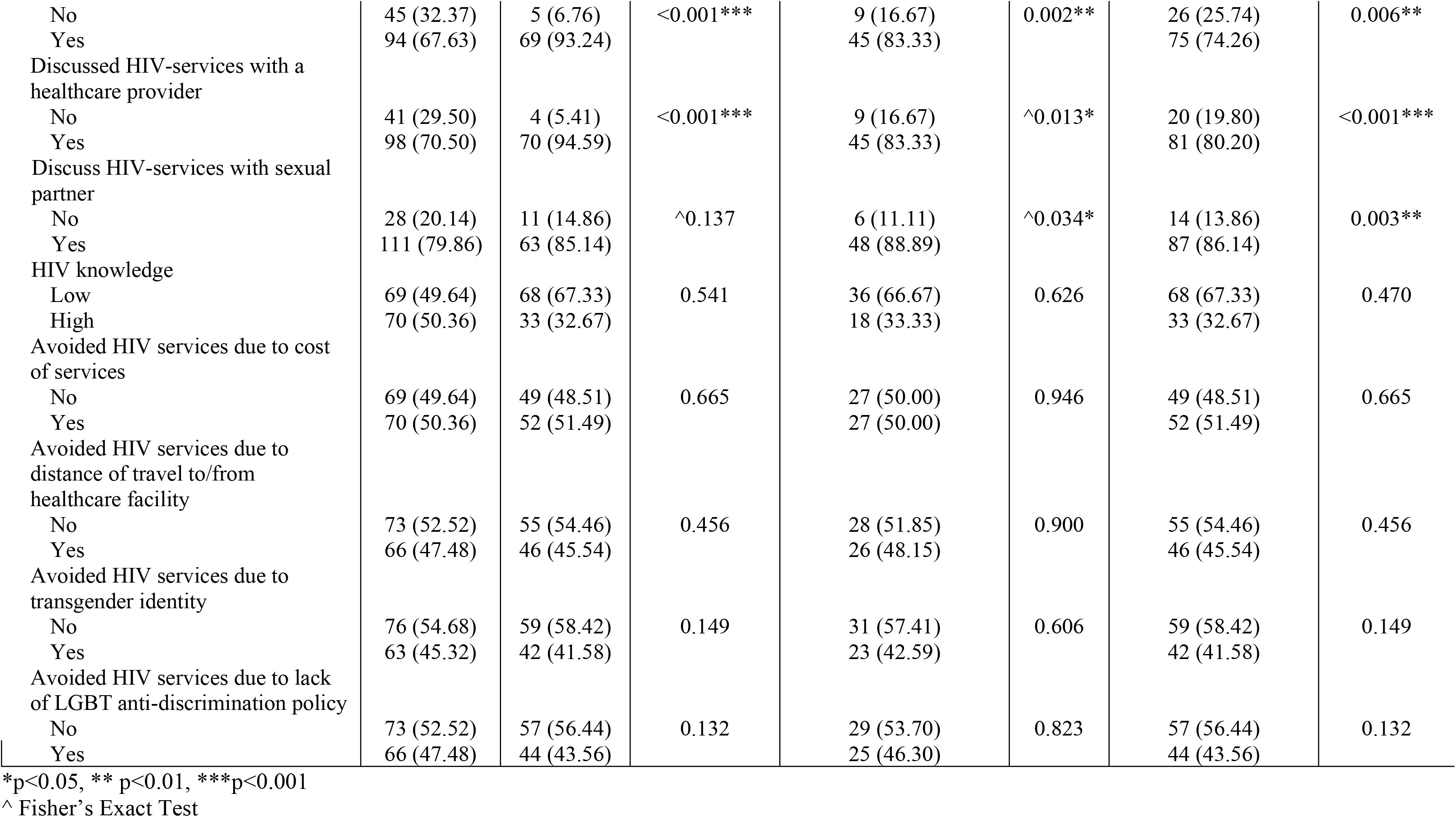
Study sample characteristics of Filipina transfeminine adults (n=139).

|  | Total | <u>PrEP Awareness</u> |  | <u>PrEP Discussion among Trans Friends</u> |  | <u>Interest in Long-Acting Injectable PrEP</u> |  |
| --- | --- | --- | --- | --- | --- | --- | --- |
| | n (%) | Yes n (%) | $\chi^2$ test, p-values | Yes n (%) | $\chi^2$ test, p-values | Very Interested n (%) | $\chi^2$ test, p-values |
| Total | 139 | 74 (53.24) |  | 54 (38.85) |  | 101 (72.66) |  |
| **Demographics** |  |  |  |  |  |  |  |
| Age |  |  |  |  |  |  |  |
| 18 to 24 | 45 (32.37) | 19 (25.68) | $\wedge 0.108$ | 9 (16.67) | $\wedge 0.014^*$ | 35 (36.46) | $\wedge 0.255$ |
| 25 to 29 | 54 (38.85) | 29 (39.19) |  | 27 (50.00) |  | 37 (38.54) |  |
| 30 to 34 | 20 (14.39) | 15 (20.27) |  | 9 (16.67) |  | 16 (16.67) |  |
| 35 years or more | 20 (14.39) | 11 (14.86) |  | 9 (16.67) |  | 8 (8.33) |  |
| Current living location |  |  |  |  |  |  |  |
| Metropolitan Manila | 110 (79.14) | 56 (75.68) | $\wedge 0.304$ | 40 (74.07) | 0.242 | 75 (74.26) | $\wedge 0.020^*$ |
| Central Visayas | 29 (20.86) | 18 (24.32) |  | 14 (25.93) |  | 26 (25.74) |  |
| Highest educational attainment |  |  |  |  |  |  |  |
| High School or below | 55 (39.57) | 22 (29.73) | $\wedge 0.041^*$ | 20 (37.04) | $\wedge 0.920$ | 36 (35.64) | $\wedge 0.319$ |
| Some College | 23 (16.55) | 15 (20.27) |  | 9 (16.67) |  | 18 (17.82) |  |
| College or beyond | 61 (43.88) | 37 (50.00) |  | 25 (46.30) |  | 47 (46.53) |  |
| Past year income (Philippine Pesos) |  |  |  |  |  |  |  |
| No Income / Less than ₱10,000 | 77 (55.40) | 31 (41.89) | $\wedge 0.006^{**}$ | 27 (50.00) | $\wedge 0.346$ | 50 (49.50) | $\wedge 0.044^*$ |
| ₱10,000 – less than P20,000 | 27 (19.42) | 17 (22.97) |  | 11 (20.37) |  | 24 (23.76) |  |
| ₱20,000 – less than P30,000 | 11 (7.91) | 8 (10.81) |  | 7 (12.96) |  | 7 (6.93) |  |
| ₱30,000+ | 24 (17.27) | 18 (24.32) |  | 9 (16.67) |  | 20 (19.80) |  |
| Religious affiliation |  |  |  |  |  |  |  |
| Catholic | 110 (79.14) | 50 (67.57) | $\wedge < 0.001^{***}$ | 41 (75.93) | $\wedge 0.404$ | 79 (78.22) | $\wedge 0.545$ |
| Non-Catholic (e.g., Protestant, Christian) | 21 (15.11) | 19 (25.68) |  | 8 (14.81) |  | 17 (16.83) |  |
| Non-religious | 8 (5.76) | 5 (6.76) |  | 5 (9.26) |  | 5 (4.95) |  |
| Sexual orientation |  |  |  |  |  |  |  |
| Gay / Lesbian | 77 (55.40) | 41 (55.41) | $\wedge 0.866$ | 34 (62.96) | $\wedge 0.270$ | 54 (53.47) | $\wedge 0.891$ |
| Bisexual | 18 (12.95) | 10 (13.51) |  | 5 (9.26) |  | 14 (13.86) |  |
| Straight | 26 (18.71) | 15 (20.27) |  | 11 (20.37) |  | 20 (19.80) |  |
| Another sexual orientation | 18 (12.95) | 8 (10.81) |  | 4 (7.41) |  | 13 (12.87) |  |
| **Social Marginalization Factors** |  |  |  |  |  |  |  |
| Ever homelessness |  |  |  |  |  |  |  |
| No | 101 (73.19) | 49 (67.12) | ^0.123 | 36 (67.92) | 0.324 | 71 (71.00) | ^0.396 |
| Yes | 37 (26.81) | 24 (32.88) |  | 17 (32.08) |  | 29 (29.00) |  |
| Currently unemployed |  |  |  |  |  |  |  |
| No | 59 (42.45) | 53 (71.62) | ^<0.001** | 33 (61.11) | 0.598 | 39 (38.61) | ^0.178 |
| Yes | 80 (57.55) | 21 (28.38) |  | 21 (38.89) |  | 62 (61.39) |  |
| Recent (<4 mos) sex work engagement |  |  |  |  |  |  |  |
| No | 67 (48.20) | 41 (55.41) | ^0.089 | 23 (42.59) | 0.303 | 46 (45.54) | ^0.344 |
| Yes | 72 (51.80) | 33 (44.59) |  | 31 (57.41) |  | 55 (54.46) |  |
| **Community-level Factors** |  |  |  |  |  |  |  |
| Social cohesion |  |  |  |  |  |  |  |
| Low | 66 (47.48) | 23 (31.08) | ^0.010** | 16 (29.63) | ^0.023* | 37 (36.63) | 0.047 |
| High | 73 (52.52) | 51 (68.92) |  | 38 (70.37) |  | 64 (63.37) |  |
| General social participation |  |  |  |  |  |  |  |
| Low | 86 (61.87) | 39 (52.70) | ^0.023* | 29 (53.70) | 0.114 | 62 (61.39) | 0.848 |
| High | 53 (38.13) | 35 (47.30) |  | 25 (46.30) |  | 39 (38.61) |  |
| LGBT-specific social participation |  |  |  |  |  |  |  |
| Low | 60 (43.17) | 20 (27.03) | ^<0.001*** | 16 (29.63) | 0.010* | 36 (35.64) | 0.004** |
| High | 79 (56.83) | 54 (72.97) |  | 38 (70.37) |  | 65 (64.36) |  |
| Healthcare Accessibility |  |  |  |  |  |  |  |
| Poor/Fair | 74 (53.24) | 37 (50.00) | ^0.025* | 26 (48.15) | 0.038* | 55 (54.46) | 0.085 |
| Good/Excellent | 65 (46.76) | 37 (50.00) |  | 28 (51.85) |  | 46 (45.54) |  |
| **HIV and Other Healthcare Indicators** |  |  |  |  |  |  |  |
| Current health insurance |  |  |  |  |  |  |  |
| No | 94 (67.63) | 40 (54.05) | ^<0.001*** | 29 (53.70) | 0.005** | 62 (61.39) | ^0.014* |
| Yes | 45 (32.37) | 34 (45.95) |  | 25 (46.30) |  | 39 (38.61) |  |
| Ever taken hormones for gender affirmation |  |  |  |  |  |  |  |
| No | 49 (35.25) | 24 (32.43) | 0.458 | 13 (24.07) | 0.028* | 31 (30.69) | 0.076 |
| Yes | 90 (64.75) | 50 (67.57) |  | 41 (75.93) |  | 70 (69.31) |  |
| Ever had surgery for gender affirmation |  |  |  |  |  |  |  |
| No | 112 (80.58) | 58 (78.38) | 0.485 | 42 (77.78) | 0.506 | 79 (78.22) | 0.338 |
| Yes | 27 (19.42) | 16 (21.62) |  | 12 (22.22) |  | 22 (21.78) |  |
| Healthcare discrimination due to transgender identity |  |  |  |  |  |  |  |
| No | 77 (55.40) | 37 (50.00) | 0.172 | 21 (38.89) | 0.003** | 55 (54.46) | 0.716 |
| Yes | 62 (44.60) | 37 (50.00) |  | 33 (61.11) |  | 46 (45.54) |  |
| Ever had HIV test |  |  |  |  |  |  |  |
| No | 45 (32.37) | 5 (6.76) | <0.001*** | 9 (16.67) | 0.002** | 26 (25.74) | 0.006** |
| Yes | 94 (67.63) | 69 (93.24) |  | 45 (83.33) |  | 75 (74.26) |  |
| Discussed HIV-services with a healthcare provider |  |  |  |  |  |  |  |
| No | 41 (29.50) | 4 (5.41) | <0.001*** | 9 (16.67) | ^0.013* | 20 (19.80) | <0.001*** |
| Yes | 98 (70.50) | 70 (94.59) |  | 45 (83.33) |  | 81 (80.20) |  |
| Discuss HIV-services with sexual partner |  |  |  |  |  |  |  |
| No | 28 (20.14) | 11 (14.86) | ^0.137 | 6 (11.11) | ^0.034* | 14 (13.86) | 0.003** |
| Yes | 111 (79.86) | 63 (85.14) |  | 48 (88.89) |  | 87 (86.14) |  |
| HIV knowledge |  |  |  |  |  |  |  |
| Low | 69 (49.64) | 68 (67.33) | 0.541 | 36 (66.67) | 0.626 | 68 (67.33) | 0.470 |
| High | 70 (50.36) | 33 (32.67) |  | 18 (33.33) |  | 33 (32.67) |  |
| Avoided HIV services due to cost of services |  |  |  |  |  |  |  |
| No | 69 (49.64) | 49 (48.51) | 0.665 | 27 (50.00) | 0.946 | 49 (48.51) | 0.665 |
| Yes | 70 (50.36) | 52 (51.49) |  | 27 (50.00) |  | 52 (51.49) |  |
| Avoided HIV services due to distance of travel to/from healthcare facility |  |  |  |  |  |  |  |
| No | 73 (52.52) | 55 (54.46) | 0.456 | 28 (51.85) | 0.900 | 55 (54.46) | 0.456 |
| Yes | 66 (47.48) | 46 (45.54) |  | 26 (48.15) |  | 46 (45.54) |  |
| Avoided HIV services due to transgender identity |  |  |  |  |  |  |  |
| No | 76 (54.68) | 59 (58.42) | 0.149 | 31 (57.41) | 0.606 | 59 (58.42) | 0.149 |
| Yes | 63 (45.32) | 42 (41.58) |  | 23 (42.59) |  | 42 (41.58) |  |
| Avoided HIV services due to lack of LGBT anti-discrimination policy |  |  |  |  |  |  |  |
| No | 73 (52.52) | 57 (56.44) | 0.132 | 29 (53.70) | 0.823 | 57 (56.44) | 0.132 |
| Yes | 66 (47.48) | 44 (43.56) |  | 25 (46.30) |  | 44 (43.56) |  |

\*p<0.05, \*\* p<0.01, \*\*\*p<0.001
^ Fisher's Exact Test

The majority of the respondents had high social cohesion and high LGBT-specific social participation (53% and 57%, respectively). Less than half of the respondents had high general social participation and good/excellent healthcare accessibility (38% and 47%, respectively). Additionally, 32% of respondents had current health insurance, 65% had ever taken gender-affirming hormones, 19% had gender-affirming surgery, and 45% faced healthcare discrimination due to their transgender identity. Most respondents had previously received an HIV test (68%), discussed HIV with their provider (71%) as well as their sexual partner (80%), and displayed high HIV knowledge (50%). About half of the respondents experienced avoidance of HIV services due to: cost (50%), distance to/from a facility (47%), their transgender identity (45%), and lack of LGBT anti-discrimination policy (47%).

### PrEP Outcomes

As shown in S1 Fig 1, 53% of the sample were aware of PrEP, 39% had discussed PrEP with their trans friends, and 73% were interested in LAI-PrEP.

### Multivariable logistic regression analyses of PrEP Outcomes

Table 2 shows the final adjusted multivariable logistic regression models with variables selected by lasso procedure per PrEP outcome. Specifically, the first adjusted model (outcome: PrEP awareness) included the following lasso-selected factors: age, past year income, religious affiliation, ever homelessness, current unemployment, general social participation, current health insurance, healthcare discrimination due to transgender identity, ever had HIV test, discussed HIV-services with a healthcare provider, and HIV knowledge. The second adjusted model (outcome: PrEP Discussion among trans friends) included the following lasso-selected factors: age, sexual orientation, recent sex work engagement, LGBT-specific social participation, healthcare accessibility, current health insurance, ever taken hormones for gender affirmation, ever had HIV test, discussed HIV-services with a healthcare provider and with a sexual partner. And lastly, the third adjusted model (outcome: interest in LAI-PrEP) the following lasso-selected factors: currently living location, discussed HIV services with a healthcare provider, and with a sexual partner.

**Table 2.**
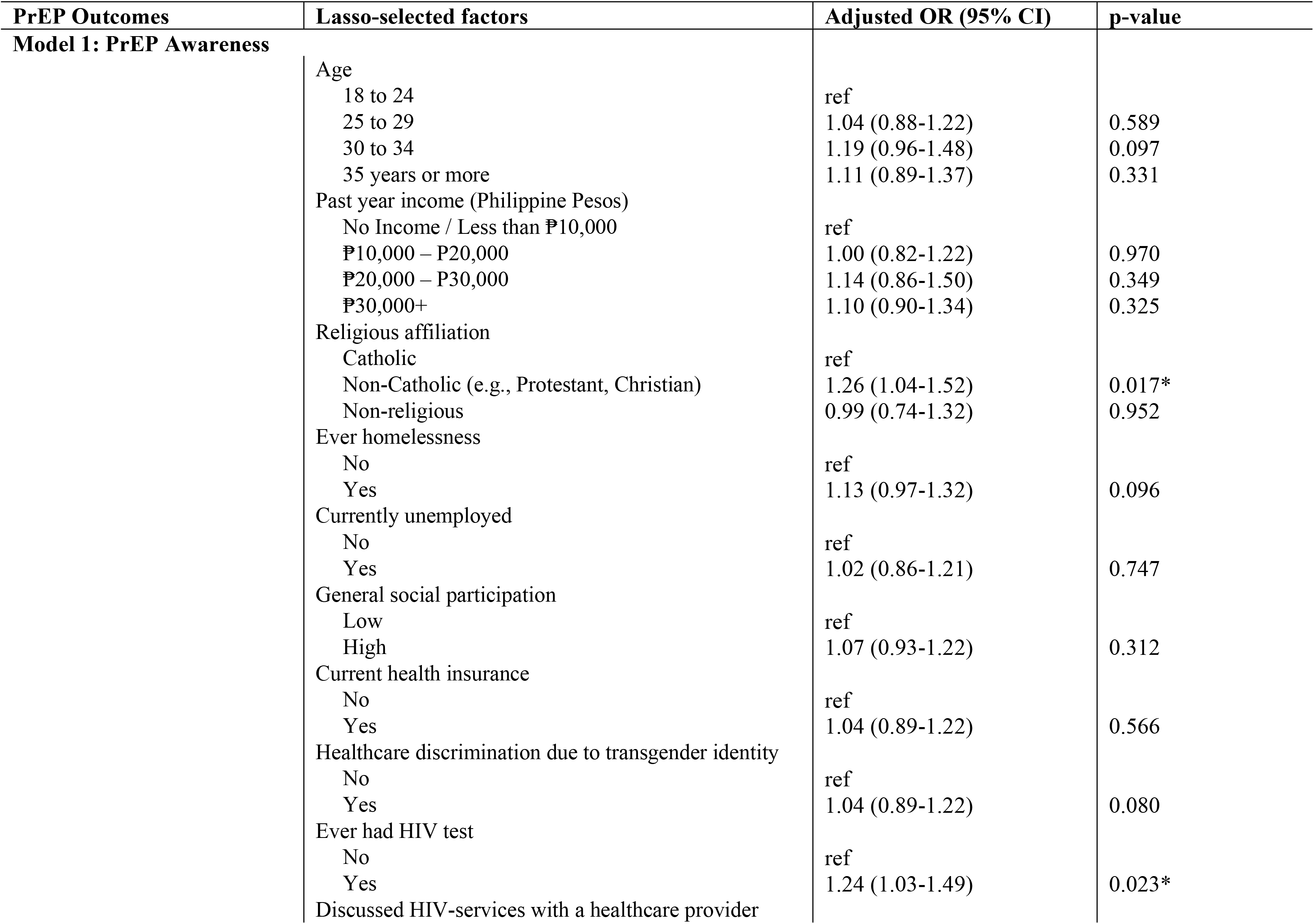
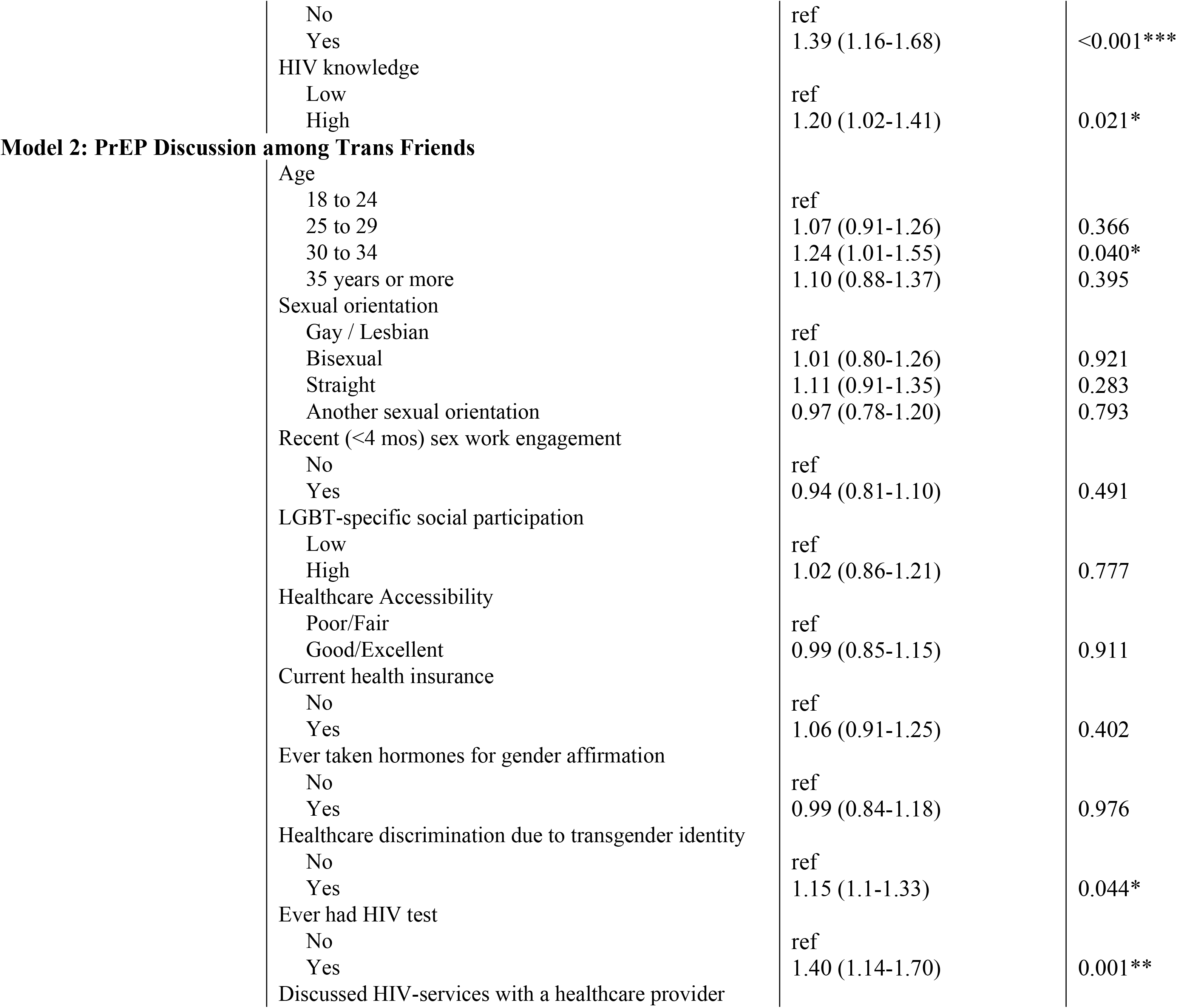
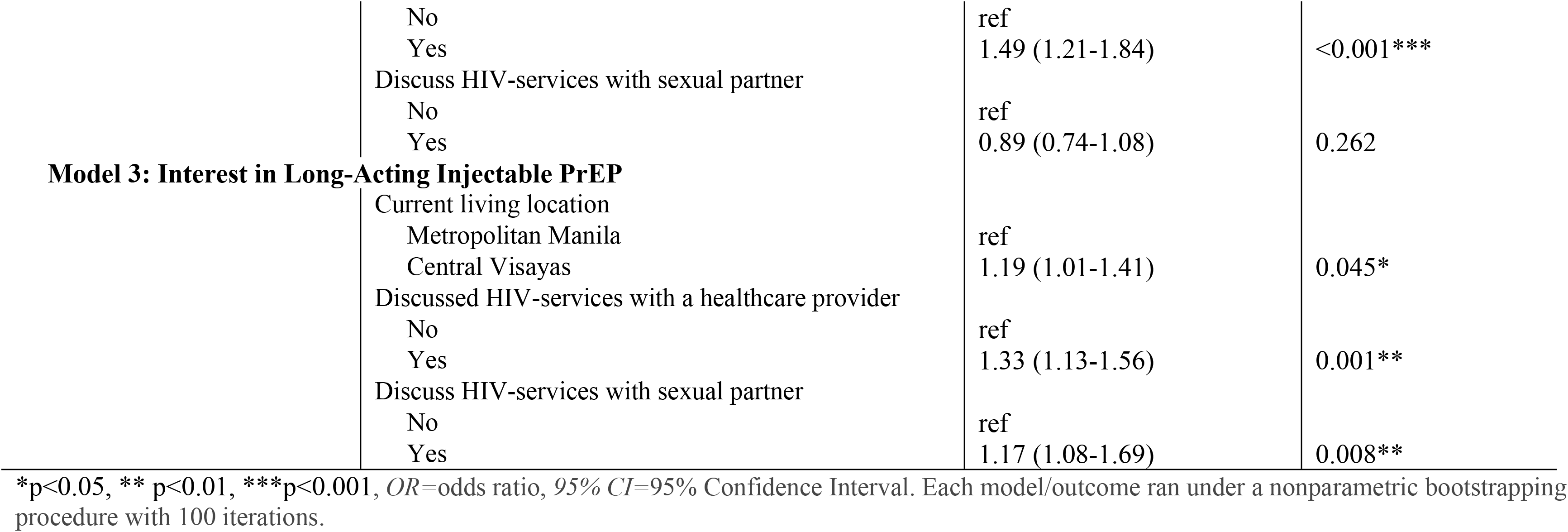
Results of adjusted multivariable logistic regression with lasso variable selection: Lasso-selected factors associated with PrEP awareness, PrEP discussion among trans friends, and interest in LAI-PrEP.

In the final model of PrEP awareness (Model 1), higher odds of PrEP awareness were associated with respondents who were non-Catholic (adjusted odds ratio [aOR]=1.26, 95% Confidence Interval [CI]=1.04-1.52), had an HIV test (aOR=1.24, 95%CI=1.03-1.49), had discussed HIV services with a healthcare provider (aOR=1.39, 95%CI=1.16-1.68), and had high HIV knowledge (aOR=1.20, 95%CI=1.02-1.41).

Moreover, higher odds of PrEP discussion among trans friends (Model 2) were associated with respondents who were between 30-34 years older (compared to 18-24 years old) (aOR=1.24, 95%CI=1.01-1.55), had experienced healthcare discrimination due to transgender identity (aOR=1.15, 95%CI=1.1-1.33), had an HIV test (aOR=1.40, 95%CI=1.14-1.70), and had discussed HIV services with a healthcare provider (aOR=1.49, 95%CI=1.21-1.84).

Finally, the odds of being interested in LAI-PrEP (Model 3) were significantly higher among respondents who were currently living in Central Visayas (aOR=1.19, 95%CI=1.01-1.41), had discussed HIV services with a healthcare provider (aOR=1.33, 95%CI=1.13-1.56), and had discussed HIV services with a sexual partner (aOR=1.17, 95%CI=1.08-1.69).

## Discussion

To our team’s expertise, this is the first study to examine LAI-PrEP outcomes among transfeminine adults in the Philippines. This setting is a compelling national context for understanding the implementation factors to optimize LAI-PrEP, based on the concentrated growth in HIV diagnoses among transfeminine people in the Philippines and the country’s stated commitment to providing PrEP medications to priority populations. Results from this analysis, as discussed below, provide vital insights into future LAI-PrEP studies and programming for this population.

Notably, all three PrEP outcomes were associated with having a discussion about HIV services with a healthcare provider, reflecting a critical point for future implementation of this intervention. Although the Philippines’ Health Department has articulated a commitment to providing access to PrEP mediations for key populations in the Philippines, efforts are needed to train providers in delivering inclusive care for trans populations. Previous research from the Philippines has corroborated the critical importance of non-discriminatory and gender affirmative health settings as the foundation for the delivery of HIV services to transgender people [39]. Additionally, systems are necessary to monitor for trans-discriminatory practices and hold providers and service settings accountable in instances where discrimination and stigma are reported.

Overall, PrEP awareness was relatively low with just over half of the sample having heard of PrEP. This level of awareness might reflect the very recent approval of PrEP as part of the Philippines government’s medical formulary, with a potential for higher awareness over time.

However, research in other settings has noted that PrEP messaging tends to prioritize MSM populations exclusively (e.g., in print and social media campaigns) and has called for the intentional creation of trans-specific messaging to maximize visibility and uptake [16]. Our findings also reveal patterns of PrEP awareness within this transfeminine sample. Of note, those who identified as being Catholic had lower PrEP awareness, suggesting a need to explore the possible roles of stigma and religion as contributors to HIV prevention and risk in this predominantly Catholic setting. Moreover, while many LGBTQ people in the Philippines identify as religious from one recent study [40], the extent to which religiosity and Catholic affiliation dictate sexual behavior and health-seeking behavior specific to sexual health including PrEP is unknown.

Even fewer participants (39%) had ever discussed PrEP with their friends. Social communication about PrEP has proven to be a challenge in many settings due to PrEP-related stigma in many parts of the world [8]. For example, studies have suggested some of the judgmental beliefs attributed to people interested in PrEP – e.g., beliefs that equate PrEP with promiscuity, risk-taking, shame [41, 42]. Health providers are often viewed as barriers to PrEP due to patients’ concerns about stigma when seeking advice about PrEP [42], and due to providers’ lack of both HIV knowledge and trans-specific competency [39]. Indeed, our findings indicated that prior experience of gender-based discrimination in a healthcare setting was negatively associated with PrEP discussion among trans friends. This suggests a need for interventions and support systems that counteract internalized stigma and shame, for example, based on negative health-seeking experiences, and that empower transfeminine people with comfort in openly discussing PrEP as an option for HIV prevention with their peers. These efforts are particularly needed for younger transfeminine people who might lack comfort and experience handling these discussions, as our findings show that those ages 18 to 24 were significantly less likely to discuss PrEP with their peers. Given the lack of comprehensive and LGBTQ-inclusive sex education programming in public schools in the Philippines that may otherwise socialize them to frank discussions with peers about sexuality and sexual health [43-45], it is not surprising to find that younger Filipina transfeminine adults are less to discuss PrEP with trans friends compared to older groups. This finding may also reflect the lack of capacity or comfort to articulate both broad and specific sexuality and sexual health concerns as young adults who may not have disclosed their gender identities or may be newly out. Future HIV education programs for this young adult group must take into account the minimal or total lack of formative experiences they have to discuss LGBTQ sexuality concerns with peers and the skills necessary to develop comfort in these peer-to-peer discussions. As such, one point of further research would be for future interventions to integrate developmentally appropriate and culturally-sensitive peer-led education programs that may be utilized for this subpopulation.

The majority of the sample reported interest in LAI-PREP thereby indicating a target for future intervention development around this modality. Notably, interest in LAI-PREP was more strongly endorsed by those who had previously discussed HIV services with a sexual partner, suggesting an opportunity for couples-involved strategies to promote this modality as a strategy to promote sexual safety and satisfaction within relationships. Additionally, interest was also stronger among those who were currently based outside of Metro Manila, suggesting the geographic appeal of this long-acting modality among those who might not have proximity to the nation’s HIV most resourced hospital and public health settings. Leveraging innovative strategies and lessons learned from the Philippines’ local community-based organizations’ responses to delivering HIV services in light of the recent COVID-19 pandemic could be promising to integrate into future LAI-PrEP programming [46]; this includes improving logistics and protocols such as online remote service delivery (e.g., counseling, referrals, refill appointments), a continuance of supply chain of medications, and app-based strategies to reach individuals who may be remote or experience travel difficulties to clinics.

While transfeminine adults in our sample showed high interest in taking LAI-PrEP, it is necessary for future studies aiming to inform PrEP implementation strategies to ascertain what transfeminine adults know about LAI-PrEP, including attitudes and beliefs about LAI-PrEP and how to best deliver it to their communities and social groups [8]. This includes understanding key health communication strategies such as efficacy messaging and framings about LAI-PrEP that resonate with other health priorities among communities of transfeminine adults such as access to hormones and gender-inclusive providers [47]. Future research priorities also include examining preferences for how to best market and package LAI-PrEP with gender-affirming care, along with exploration of other PrEP options and regimens such as daily PrEP, intermittent (2-1-1) dosing, and implants [47, 48] – to ensure that LAI-PrEP fully aligns with and directly benefits Filipina transfeminine adults’ goals for HIV prevention and gender-affirmation [49].

Given that our study only examined three PrEP outcomes (awareness, discussion among friends, and interest), future studies must also expand the scope of examination to other outcomes along the PrEP continua (e.g., awareness, uptake, adherence, and retention) [29], to fully optimize, track, and evaluate the progress of LAI-PrEP implementation programs as it gets delivered at a population-level in the Philippines. This includes formatively understanding and conceptualizing if there are any other unique points or steps to the care continuum that are unique to LAI-PrEP among transfeminine adults.

Lastly, while findings from adjusted models specific to social marginalization such as homelessness, unemployment, and recent engagement in sex work were not significantly predictive of PrEP outcomes, it is noteworthy for future studies to explore and investigate whether such subpopulations have unique needs specific to LAI-PrEP continuum that may not have been captured in this study. Given the well-recognized role of social marginalization in predisposing communities to higher HIV risks and lower engagement in HIV services [50-52], it is vital for future implementation research to qualitatively explore how LAI-PrEP may be accepted by transfeminine individuals who experience and/or are placed in such socially marginalized settings.

### Limitations

There are several limitations to this study. First, this study is limited in its transferability to other settings in the Philippines due to our recruitment focus on the two largest metropolitan settings in the country. Specifically, in addition to our convenient online sampling, we only recruited in two sites where the HIV epidemic is highly prevalent and only included transfeminine adults. As such, our sample may not capture the diversity of trans communities in the Philippines, particularly those who do not have access to the internet, are not living in the study’s recruited areas, or transmasculine and nonbinary adults who are also part of trans communities and are impacted by the epidemic [53]. Second, due to the cross-sectional nature of this study’s data collection design, we are unable to ascertain temporal patterns with regard to PrEP outcomes. Lastly, given that our study utilized secondary analysis from an existing dataset, we were unable incorporate additional detailed PrEP-specific outcomes. Future research is necessary to explore socio-ecological factors that are vital in LAI-PrEP implementation in this population and setting.

## Conclusion

This study provides important foundational work for understanding how LAI-PrEP may fit into the appropriate HIV prevention infrastructure among Filipina transfeminine adults. Though we categorize initial interest in LAI-PrEP, critical other insights will be needed to effectively build towards LAI-PrEP rollout among Filipina transfeminine adults. Building upon these nascent findings, we encourage future work on these fronts, especially moving beyond just research and policy goals and ensuring that other efforts actively engage with community-based organization and their aims and needs. In doing so, the Philippines’ HIV response can continue to build towards improvements in equity among transfeminine adults and other impact subpopulations, an important and essential goal.

## Data Availability

Given that this study contains data with potentially sensitive information, data from this study are available upon request. Contact the Brown University Human Research Protection Program Institutional Review Committee for data requests.

## Acknowledgments

We are grateful for all of the Filipina transfeminine participants who took part in this study. This study was supported by the following sponsors: National Institute on Drug Abuse (R36DA048682), the Fogarty International Center (D43TW010565), Providence/Boston Center for AIDS Research (P30AI042853), and the National Institute of Mental Health (R21TW012010). Dr. Restar is a Robert Wood Johnson Foundation Health Policy Research Scholar and a Public Policy Fellow at amFAR, the Foundation for AIDS Research. Drs. Restar, Flores, and Wickersham are supported by the Research Education Institute for Diverse Scholars (REIDS), funded by the National Institute of Mental Health (R25MH087217). This article does not represent the official views of the sponsors.

## Compliance of Ethical Standards

### Conflict of Interest

The authors declare that they have no conflict of interest.

### Ethical approval

All procedures performed in studies involving human participants were in accordance with the ethical standards of the institutional research committee (Brown University Ethics Review Board, IRB Protocol, #1802001982) and with the 1964 Helsinki declaration and its later amendments or comparable ethical standards.

### Informed consent

Electronic informed consent was obtained from all individual participants included in the study.

---

**S1 Fig. Figure 1.**
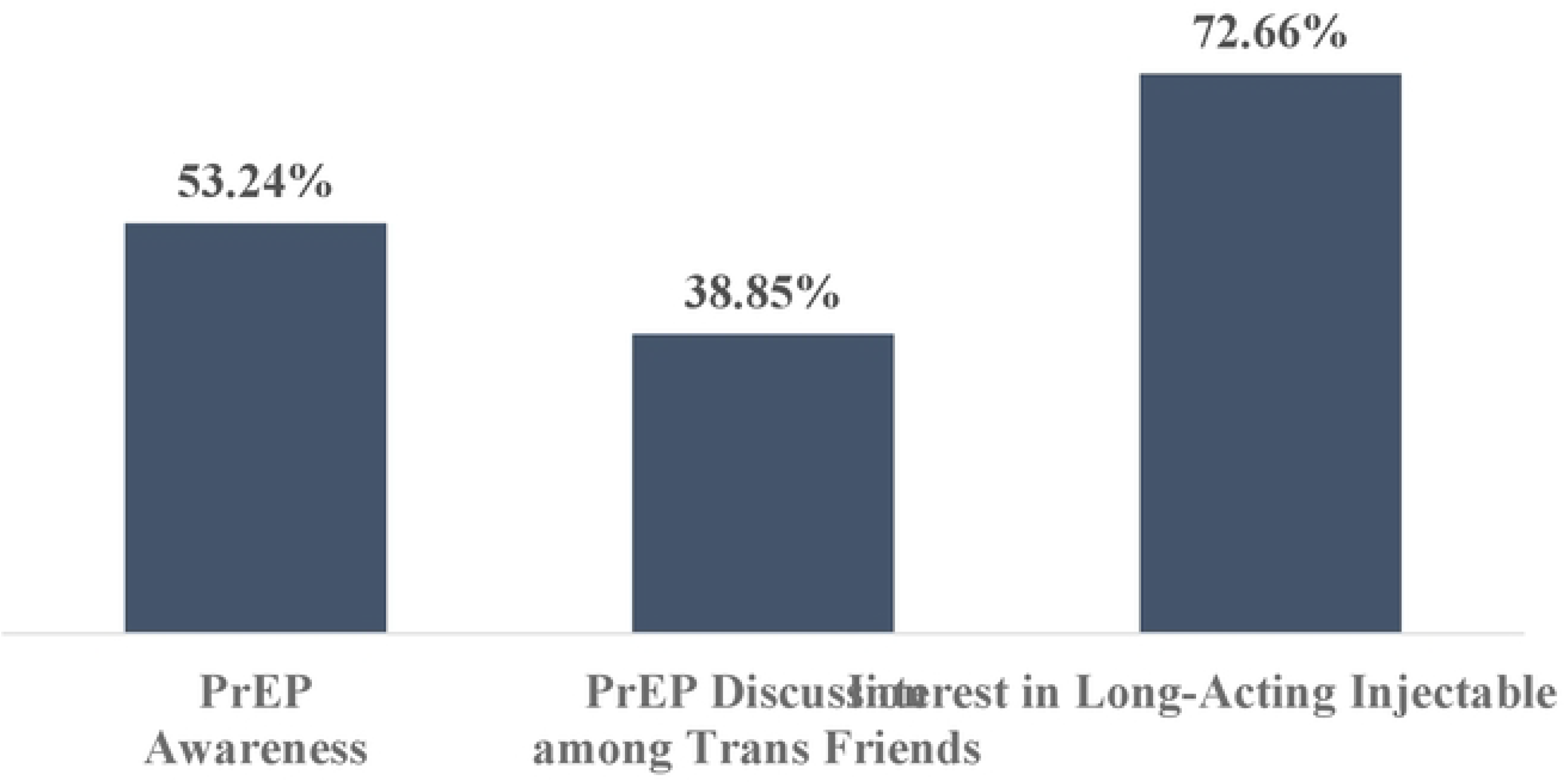
PrEP awareness, PrEP discussion among trans friends, and interest in long-acting PrEP among Filipina transfeminine adults (n=l39).

## Notes

### Competing Interest Statement

The authors have declared no competing interest.

### Funding Statement

The authors declare that they have no conflict of interest, including financial.

### Author Declarations

This study acquired approval from Institutional Review Board of the Brown University Brown University Human Research Protection Program (IRB Protocol, #1802001982).

